# Evaluating the effects of second-dose vaccine-delay policies in European countries: A simulation study based on data from Greece

**DOI:** 10.1101/2021.05.19.21257486

**Authors:** P. Barmpounakis, N. Demiris, I. Kontoyiannis, G. Pavlakis, V. Sypsa

**Affiliations:** Department of Statistics, Athens University of Economics and Business, Athens, Greece; Department of Pure Mathematics and Mathematical Statistics, University of Cambridge, Cambridge, United Kingdom; Human Retrovirus Section, National Cancer Institute, Frederick, Maryland, United States of America; Dept. of Hygiene, Epidemiology and Medical Statistics, Medical School, National and Kapodistrian University of Athens, Athens, Greece

**Author notes:** Corresponding author (PB).

## Abstract

The results of a simulation-based evaluation of several policies for vaccine rollout are reported, particularly focusing on the effects of delaying the second dose of two-dose vaccines. In the presence of limited vaccine supply, the specific policy choice is a pressing issue for several countries worldwide, and the adopted course of action will affect the extension or easing of non-pharmaceutical interventions in the next months. We employ a suitably generalised, age-structure, stochastic SEIR (Susceptible → Exposed → Infectious → Removed) epidemic model that can accommodate quantitative descriptions of the major effects resulting from distinct vaccination strategies. The different rates of social contacts among distinct age-groups (as well as some other model parameters) are informed by a recent survey conducted in Greece, but the conclusions are much more widely applicable. The results are summarised and evaluated in terms of the total number of deaths and infections as well as life years lost. The optimal strategy is found to be one based on fully vaccinating the elderly/at risk as quickly as possible, while extending the time-interval between the two vaccine doses to 12 weeks for all individuals below 75 years old, in agreement with epidemic theory which suggests targeting a combination of susceptibility and infectivity. This policy, which is similar to the approaches adopted in the UK and in Canada, is found to be effective in reducing deaths and life years lost in the period while vaccination is still being carried out.

## Introduction

Since December 2019, COVID-19 has presented a global threat to public health and to the worldwide economy, and it will likely continue to disrupt livelihoods until a high percentage of the population is vaccinated. High vaccination rates will be necessary to reach herd immunity in a short period of time. Standard theory [1] suggests that a proportion approximately equal to 1-1/*R*_0_ of the population will have to become immune (either through vaccination or previous infection) in order to effectively suppress disease transmission, where *R*_0_ is the virus’ basic reproduction number. The actual vaccination coverage required is likely to vary due to population heterogeneity, previous levels of spread of infection, and other local factors. In addition, the exact value of *R*_0_ for SARS-CoV-2 under “normal” conditions remains quite uncertain since there has been very little disease spread without some mitigation effort due to non-pharmaceutical interventions, and also due to the appearance of new variants. Therefore, constrained scenarios are likely to give a realistic estimate of the effect of distinct vaccination policies and this approach is adopted in the present paper.

Assuming a vaccination coverage between 60%-80% of the population, 3.1-4.1 billion people worldwide will need to be vaccinated [2]. With several seemingly highly efficacious vaccines available (efficacy estimated at 94.1%, 95% and 62% for Moderna, Pfizer-BioNTech and Oxford-AstraZeneca respectively) against COVID-19 disease [3-5] it appears that a return to near-normality for society and for the economy may soon be possible. Unfortunately, limited supply is currently an impediment to achieving high vaccination coverage rapidly [6].

In addition to social distancing [7] and mass testing [8], the fair allocation of scarce medical interventions such as vaccines presents ethical challenges as there are different allocation principles – treating people equally, favouring the worst-off, maximising total benefits, and promoting and rewarding social usefulness – and no single principle can address all morally relevant considerations [9,10]. Modelling studies broadly agree that, when vaccine supply is limited, prioritising the elderly is a necessary strategy to reduce COVID-19 mortality, whereas the prioritisation of younger individuals would have an impact on reducing transmission [11,12]. This agrees with epidemic theory [1] which suggests that the focus for disease control should be based on a combination of targeting susceptibility and infectivity. Therefore, assuming very scarce resources, it makes sense to focus on the most vulnerable individuals in the population. On the other extreme is the presence of nearly unlimited vaccine supply, whence aiming for achieving herd immunity is straightforward. In this work we focus on the intermediate problem which many European countries are currently facing, and prioritisation of vaccines is of the essence.

Due to supply constraints, it was decided in the UK and Canada to delay the administration of the second dose of all vaccines, based on the rationale that SARS-CoV-2 vaccination offers considerable protection after the first dose and that more people could benefit [13,14]. Although this approach seems appealing, the impact of delaying the second dose is not straightforward as it depends on several parameters such as the efficacy of the first dose in time, the levels of transmission in the population, vaccination rollout, and the vaccine profile (reduction in symptoms or in symptoms and infection) [15-17]. Country-specific information on the age-distribution of the population and social mixing patterns are also necessary to obtain realistic estimates.

The main contribution of this work is the evaluation of different vaccination strategies and their potential benefits, primarily based on data from Greece, a typical country of the EU area in terms of vaccine availability and administration, with a population of around 10.8 million people. The current strategy (strategy I) is to give the second vaccine dose three weeks after the first for the Pfizer vaccine, which currently consists of the largest portion of the available vaccines in the EU. We consider an alternative policy (strategy II) where, after the vaccination of medical personnel and those over 75, a portion of the available vaccines is distributed with a three-month time interval between the two doses. The prioritization of the medical personnel and those over 75 years old is kept constant for every strategy. Our methodology examines scenarios where the two different vaccination schedules are combined in different proportions, allowing us to explore the optimal portion of the population that should be vaccinated using the extended three-month time interval between the two doses. This is something that has not been extensively explored in the literature, since earlier studies primarily focus on finding the optimal timing of the second dose, considering that the entire population will follow the same schedule [18-20]. Extending the time interval between doses to three months aims for faster partial coverage of economically active individuals, therefore offering indirect protection to a larger proportion of the population and ultimately for potentially reducing the pandemic cost to public health and the society. This is implicitly informed by aiming for a combined effect of reducing susceptibility and infectivity in the population. Different scenarios of vaccine availability and transmission rates are considered, as well as different scenarios for the acquired immunity after the first dose for strategy II. We assess our results through simulation of an age-structured stochastic SEIR (Susceptible → Exposed → Infectious → Removed) epidemic model, suitably modified to account for the number of vaccinated individuals with different protocols.

We opted for a stochastic model because the most effective way to describe the spread of a disease is stochastic, based on the specification of the probability of disease transmission between two individuals. One may incorporate additional sources of stochasticity in the length of latent and infectious periods, but it is well known that such uncertainty is immaterial in terms of its effect on the outcome of an epidemic, particularly in a large population setting such as country-level studies; see for example [21]. Since the writing of this paper, related modelling techniques using deterministic dynamics have been proposed, that use optimization techniques to infer the model parameters and find the optimal dosing schedule [18,19]. An extension of a discrete-time, deterministic susceptible-infected-recovered model was used in [22] to plan the scheduling of first and second vaccine doses, with the underlying objective of the optimization problem being the concurrent minimization of both the healthcare impact of the epidemic and of the socio-economic impact due to the implementation of non-pharmaceutical interventions. A similarly extended SEIR model was also used to measure (through simulation) whether the effect of a standard vaccination schedule for different uptake scenarios is enough to stop the epidemic without the need of non-pharmaceutical interventions [20].

## Materials and methods

### The multitype S(V)EIR model and simulation description

The model used for the simulation of different vaccination strategies is an age-structured stochastic SEIR model that accounts for different vaccinated populations and vaccinated states, termed S(V)EIR henceforth. A schematic representation of the model is given in Fig 1.

**Fig 1.**
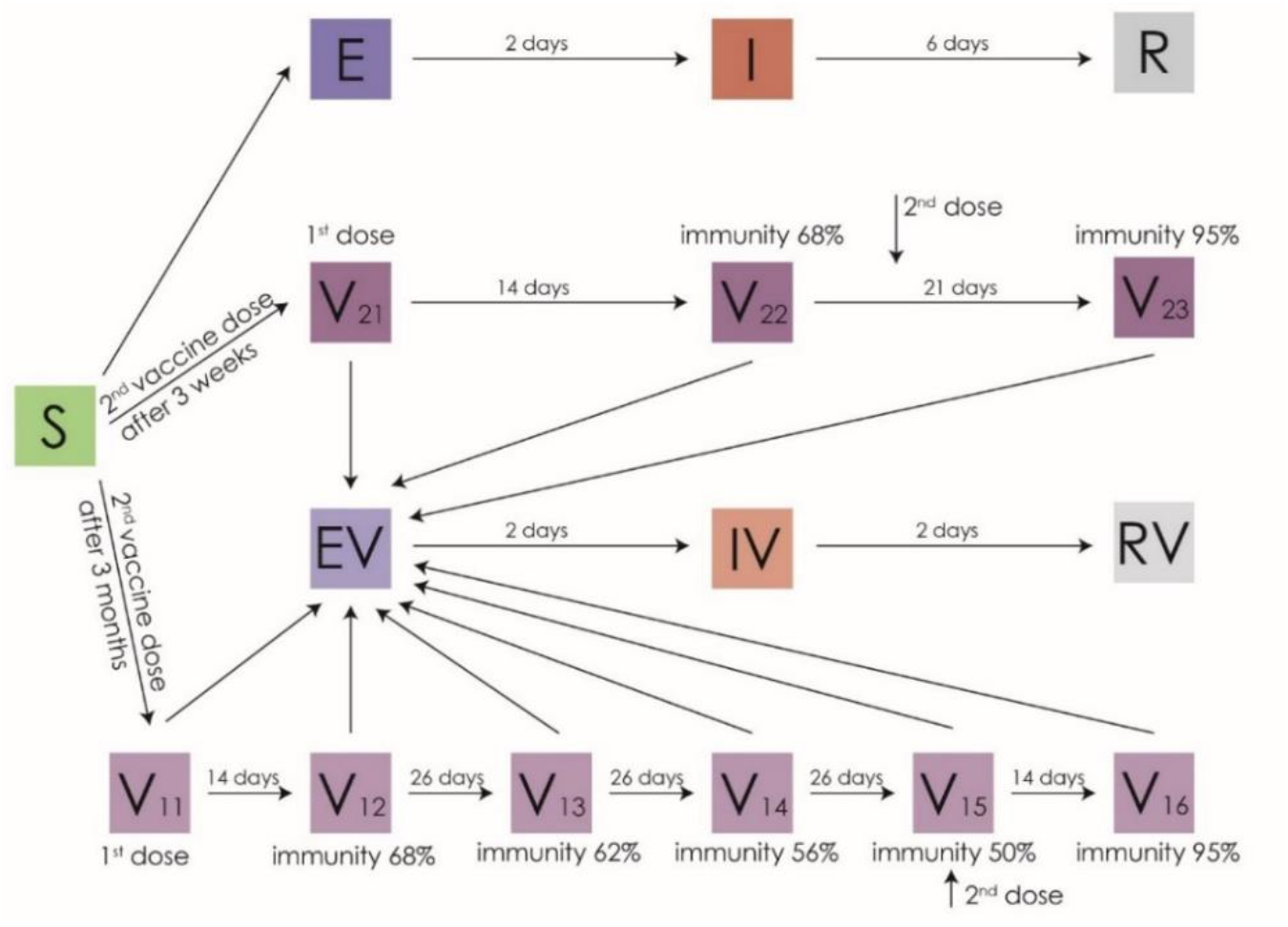
Schematic representation of the S(V)EIR epidemic model for the baseline scenario of immunity waning.

In order to evaluate the effects of different vaccination strategies, this model also accounts for the age composition of the population, the social mixing rates of different age groups, the intention to get vaccinated, as well as the different risk of death of each age group. The code for simulating the model is publicly available at: https://github.com/pbarmpounakis/Evaluating-the-effects-of-vaccine-rollout-policies-in-European-countries-A-simulation-study.

A detailed description of the model follows, while a summary of the quantitative assumptions made is given in S1 Table in Supporting information B.

### States and vaccination effect assumptions

Two groups are considered for the vaccinated people representing the two distinct vaccination categories. In vaccination group 1, individuals receive the 2^nd^ dose of the vaccine after 3 months while in vaccination group 2 it is given after 3 weeks. In both vaccination groups, individuals who received the 1^st^ dose of the vaccine move to states V11 and V21 respectively and remain fully susceptible. Two weeks after the 1^st^ dose individuals from both vaccination groups move to the second stage (V12, V22 respectively), whence immunity jumps to 68% [5]. Individuals from vaccination group 2 remain at V22 for another 7 days when they take their 2^nd^ dose and move to state 3 (V23) with their immunity jumping at 95% after two weeks. Individuals from vaccination group 1 take their second dose 78 days after entering V12 and then move to state V15 with their immunity changing based on different waning immunity scenarios described below. The stages V13, V14 and V15 account for the drop of the immunity due to waning vaccine efficacy 26, 52 and 78 days after the first dose, respectively. They move to V16 14 days later when their immunity jumps to 95%, pertaining to the reported efficacy of the mRNA-based vaccines which are mostly used in European countries [3-5]

### Transmission model assumptions

New infections from each state *s* and age group *i* follow a Binomial distribution with size given by the number of people in state *s* and age group *i*, and infection probability 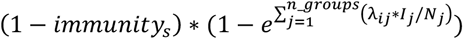, where *immunity*_*s*_ is the level of immunity at stage *s, I*_*j*_ is the number of infectious individuals at age group *j, N*_*j*_ is the total number of individuals at age group *j, λ*_*ij*_ is the *i, j* entry in the transmission matrix *λ*, and *n*_*_*_*groups* is the total number of different age groups. Following infection, individuals of age group *i* follow the Exposed (E_*i*_) → Infectious (I_*i*_) → Removed (R_*i*_) path with a constant exposure time of 2 days, based on an average incubation time of approximately 5 days [23-25] and assuming that infectiousness starts approximately 2 days prior to the occurrence of symptoms [26-28]. The infectious period is also assumed constant and set at 6 days for non-vaccinated individuals [29-31] and 2 or 3 days for vaccinated ones, depending upon the scenario of immunity waning and vaccine efficacy in reducing the infectious period [32,33]. The choices of these values are conservative assuming that in reality people experiencing influenza-like symptoms will get tested and self-isolate, resulting in lower effective infectious period. The total number of deaths is computed by multiplying the number in R_*i*_ with the infection fatality ratio (IFR) of each age group for the unvaccinated individuals as it was reported by [34] and with [IFR x 5%] for those vaccinated [35].

### Different scenarios for *R*_*t*_ and immunity

Transmission levels corresponding to a constant effective reproduction number *R*_*t*_=1.2 and *R*_*t*_=1.4 are considered along with various levels of immunity at each stage of vaccination for group 1. These choices of *R*_*t*_ suggest moderate transmission levels without the presence of a ‘hard lockdown’ for extended periods of time, closely resembling the policy that most European counties implement regarding social distancing measures during the vaccination period. *R*_*t*_ is calculated as the largest eigenvalue of the next generation matrix, using an appropriate contact matrix. The contact matrices used for the calculations of the values of *R*_*t*_ were based on a social contacts survey assessing contacts in Greece; we have used the data collected in the second half of September 2020. This contact matrix informs the relative infectivity between age-groups but, importantly, the scale is set by the value of *R*_*t*_.

We ran 1000 simulations for each scenario and computed the median as well as 90% equal-tailed uncertainty intervals. As precise data are not available for the precise course of infectivity and acquired immunity, three scenarios are considered.

#### Worst Case Scenario

It is assumed that, during the three months between the first and second dose (strategy II), the acquired immunity drops linearly to 34% (S1 Fig, Supporting information A). The effective infectious period of those vaccinated is reduced by 50% to 3 days.

#### Baseline Scenario

Here it is assumed that during the three months between the two doses (strategy II) the acquired immunity drops linearly to 50% and the infectious period of those vaccinated is set at 2 days (Fig 1).

#### Optimistic Scenario

In this case a constant immunity of 68% is assumed for the entire time between the first and second dose, and the infectious period lasts 2 days (S2 Fig, Supporting information A).

### Fraction of vaccines given to general population

Different percentages are considered for the proportion of the available vaccines distributed under strategy II. These are set to 0% (strategy I), 20%, 50% and 100% (strategy II); the resulting number of deaths and life years lost are computed in each of these cases. Moreover, in the model we assume that individuals due for the second dose have priority over those waiting to have their first dose, keeping the time interval between the two doses intact. The remaining doses available each day are given to unvaccinated individuals.

### Vaccine availability

Two levels of vaccine availability are considered; a baseline level and a limited level with reduced number of vaccines, see S1 Table in Supporting information B.

### Intention to get vaccinated

The populations’ intention to get vaccinated is informed by a telephone survey contacted by [36]; see S1 Table in Supporting Information B. We assume that after the vaccination coverage of an age group reaches the percentage of people answering “*Probably/Definitely Yes”* to whether they intend to get vaccinated, the vaccination rollout continues to the next (younger) age group. The percentage of people answering “*Probably/Definitely No”* remains unvaccinated. The individuals answering “*Don’t know/Don’t answer”* are distributed among the two other groups according to their respected adjusted percentages.

### Ethics Statement

This is a simulation study so no human participants were involved. For this reason, review and approval of this study from an ethical committee were not necessary. Since no human participants were recruited there are no available consent data or details.

Regarding the data survey based on the [36,37], since this survey was conducted by one of the authors on the current manuscript, we also provide an ethics statement for this. The protocol of the survey was approved by the Institutional Review Board of the Hellenic Scientific Committee for the Study of AIDS and STDs. Participants provided oral informed consent. The survey was conducted over the phone and participants were asked for their consent at the beginning.

## Results

Our main finding is that the optimal strategy in terms of the reduction in deaths and number of years lost, is the one that where all available vaccine doses are given under strategy II, using a time interval of three months between the two doses (Table 1).

**Table 1.** Comparison of strategy I (0% of population with 3 month interval) and strategy II (100% of population with 3 month interval)

| Strategy II vs. strategy I | Total (%) reduction under strategy II during January-December 2021 |
| --- | --- |
| Gain in number of deaths | 579 (9.04%) |
| Gain in years of life | 14802 (10.65%) |

Comparison of strategy I and strategy II (at 100% doses given) for 2021 with *R*_*t*_=1.2, under the baseline immunity scenario and standard vaccine availability. “Gain” refers to the number of *fewer* deaths and life years lost under strategy II (extended interval between doses).

The results vary between different immunity waning scenarios and different values of *R*_*t*_, but they are robust in that the optimal strategy is always found to be the one that allocates 100% of the available doses under strategy II. The intermediate allocations of 20% or 50% of the available doses for strategy II show similar mortality for the whole population with Strategy I. But because fewer younger people get infected and die, there is a reduction in total life years lost (S2-S4 Tables).

Next, we examine the resulting figures for daily deaths, life years lost, and daily infections, under the baseline immunity waning scenario, with *R*_*t*_=1.2, and with standard vaccine availability (Figs 3-5); detailed summaries of simulation results are presented in S2-S9 Tables given in Supporting information C; several additional results based on both the optimistic and worst-case scenarios are summarised in a web supplement at: https://github.com/pbarmpounakis/Evaluating-the-effects-of-vaccine-rollout-policies-in-European-countries-A-simulation-study

**Fig 2.**
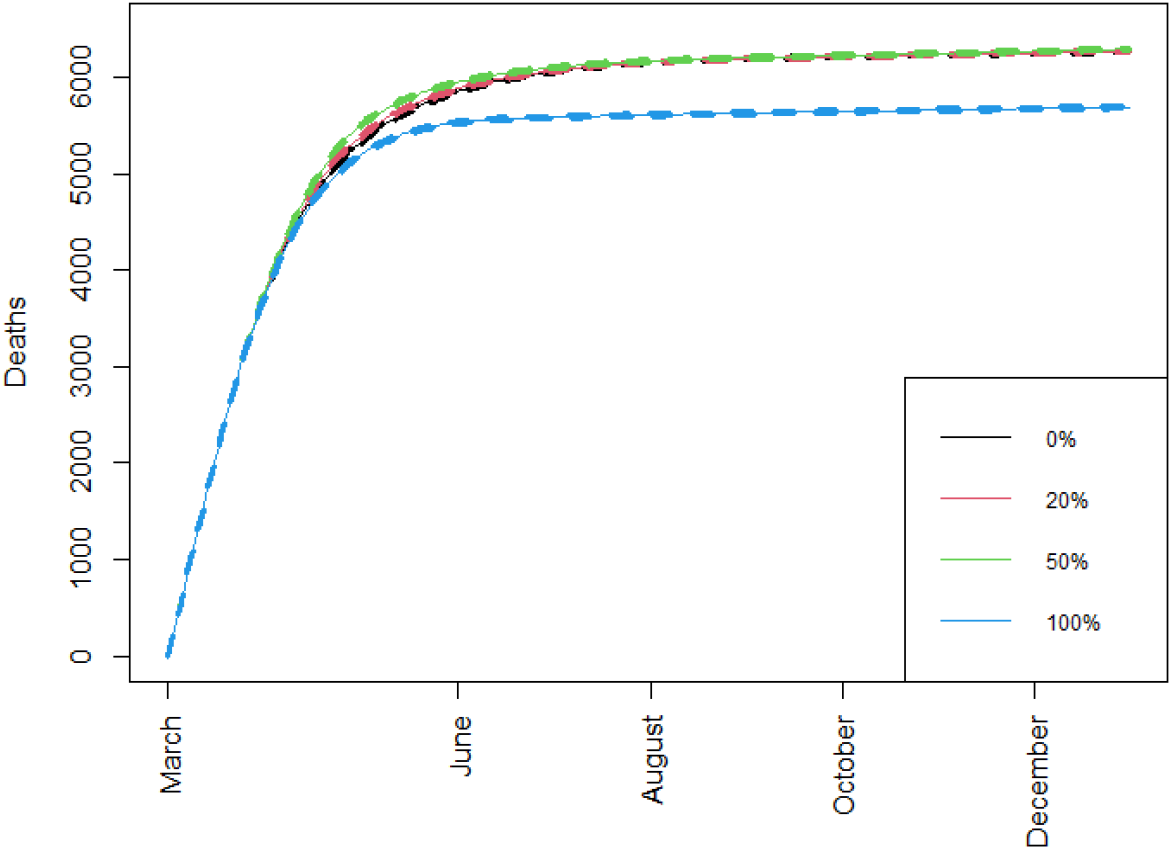
Cumulative number of deaths. Cumulative number of deaths over time when different percentages of doses are allocated under strategy II, with *Rt*=1.2, immunity drop between the two vaccine doses is at the baseline scenario, and with standard vaccine availability.

**Fig 3.**
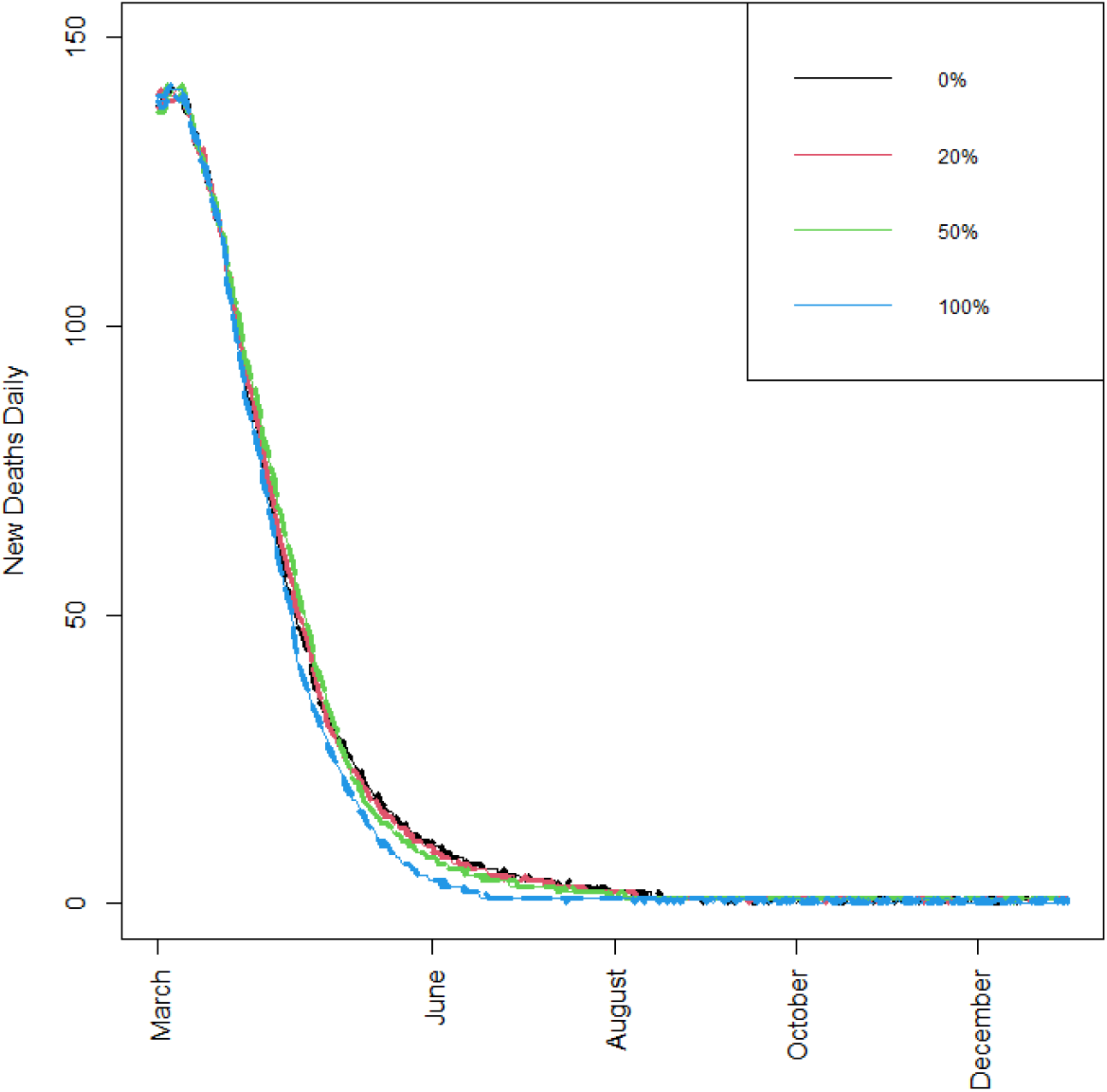
Number of new daily deaths. Number of new daily deaths, when different percentages of doses are allocated under strategy II, *Rt*=1.2, immunity drop is at the baseline scenario, and with standard vaccine availability. [A zoomed-in version of this figure is available in the supplement, offering higher resolution locally.]

**Fig 4.**
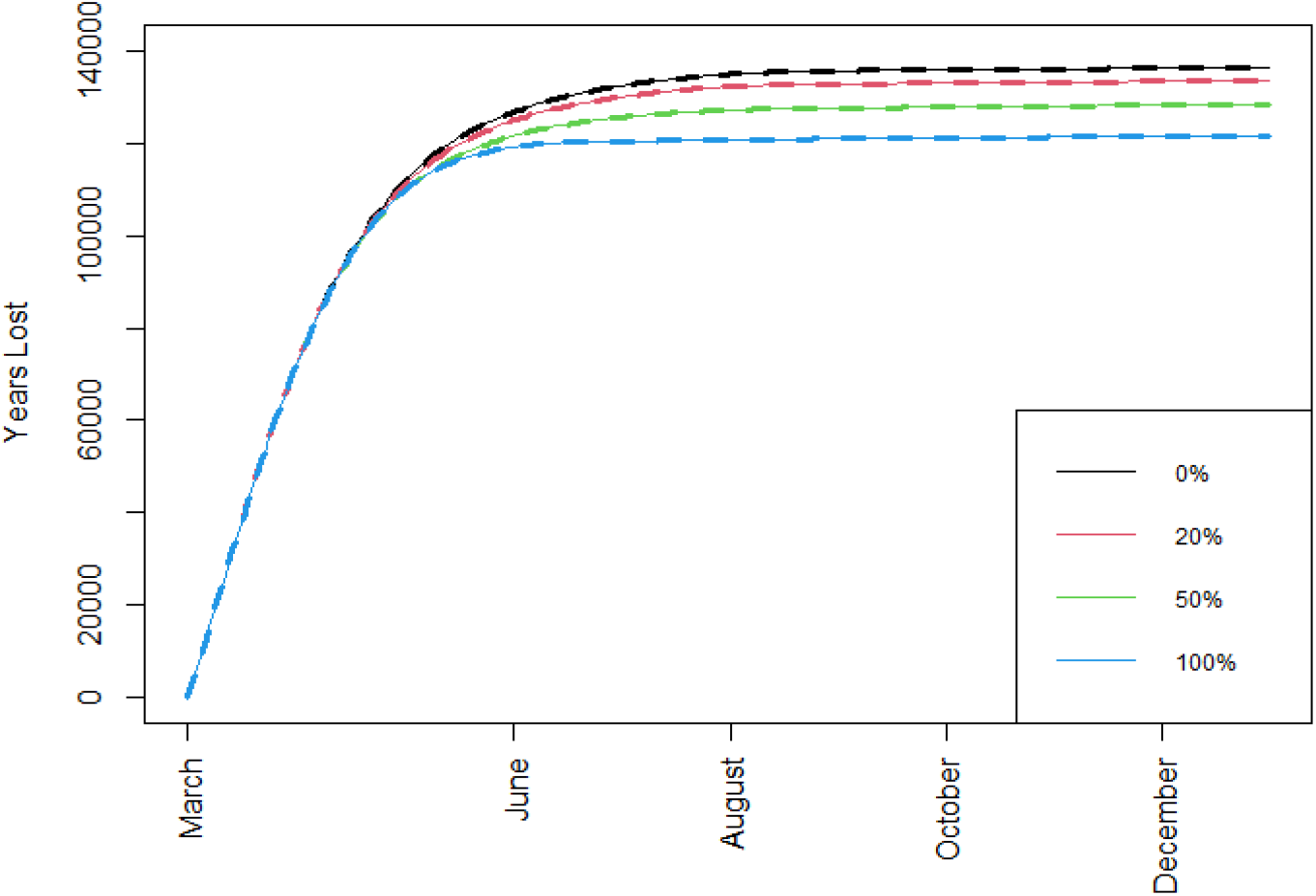
Total number of years of life lost. Total number of years of life lost when different percentages of doses are allocated under strategy II, *Rt*=1.2, immunity drop is at the baseline scenario, and with standard vaccine availability.

**Fig 5.**
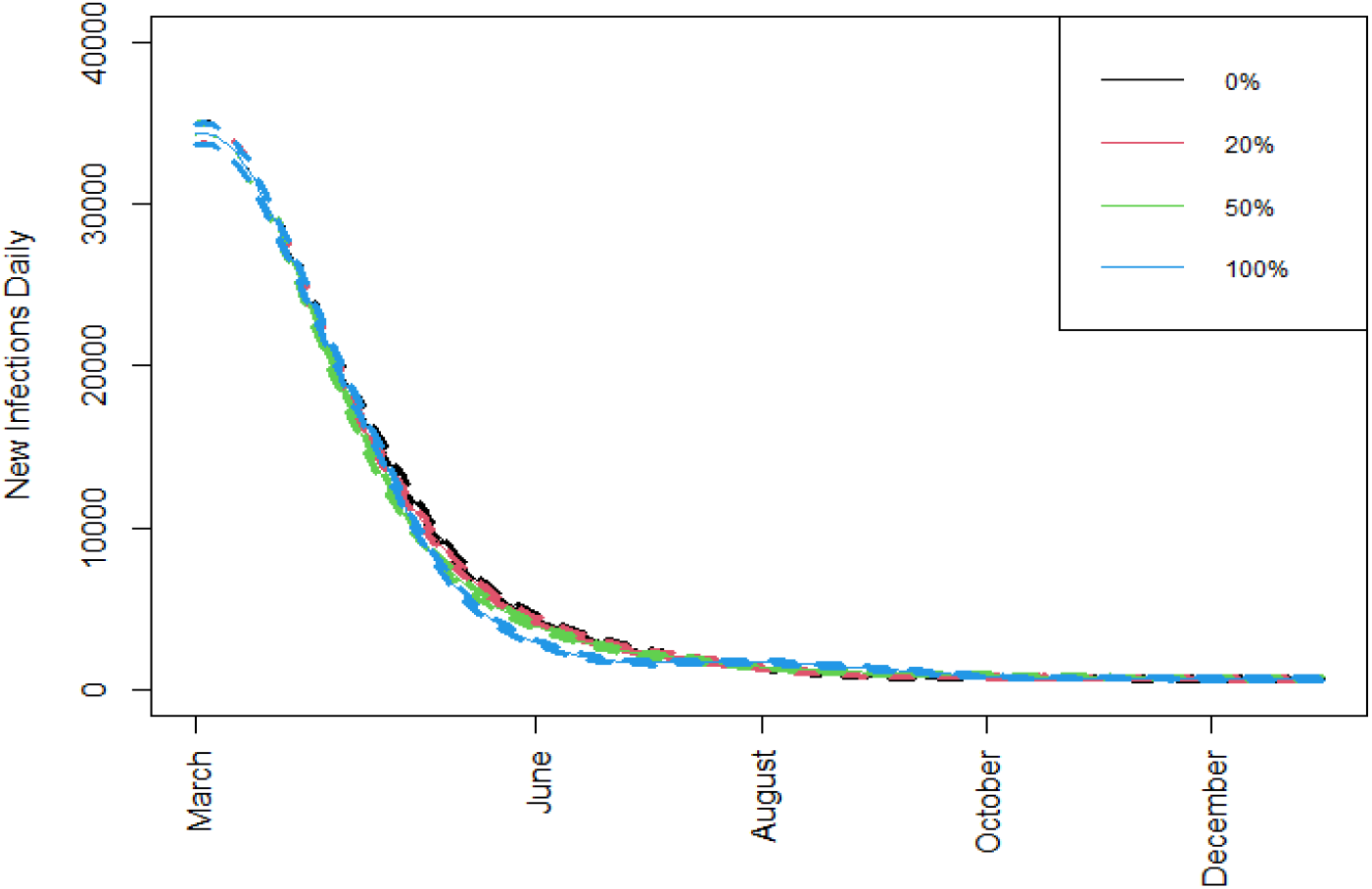
New daily infections. New daily infections when different percentages of doses are allocated under strategy II, *Rt*=1.2, immunity drop is at the baseline scenario, and with standard vaccine availability.

## Discussion

After the vaccination of medical personnel, high-risk individuals, and people aged over 75 years old with a time interval of 3-4 weeks between doses, the strategy of vaccinating the rest of the population with an interval of three months between the two doses (strategy II) can result in a significantly reduced number of deaths and years of life lost. When only 20% or 50% of the available vaccines are distributed using strategy II, the results are not significantly different to strategy I in terms of deaths, although they do provide an improvement in the number of life years saved. In conclusion, rolling out 100% of the available vaccines using the delayed second dose strategy appears to be the most effective option.

In the absence of detailed social contact data between different groups, we accounted for age groups as a surrogate for population composition, and we used the contact rate data between different age-groups from the recent survey [37]. Therefore, the results reported here offer a conservative assessment since no attempt is made to prioritize individuals with many contacts such as mass transit employees, those working in the hospitality industry, super-markets and so on. Consequently, in practice, the benefits are expected to be even greater if a more targeted approach is adopted.

We used a multitype, age-structured, stochastic epidemic model with constant transmission rate and constant exposed and infectious periods. Although this approach of course has some limitations, they are not expected to materially affect the results. First, in our model, we assumed that vaccine efficacy was mediated by a reduction in infections and not just in clinical disease. Recent modelling studies suggest that, if vaccines reduce symptomatic infection only, then the optimal protection for minimising deaths is obtained by prioritising older individuals [15]. This assumption is realistic especially in view of recent data suggesting that COVID-19 vaccines are indeed effective in the prevention of infection at least before the occurrence of the Delta strain [38,39]. Second, we assessed two scenarios for viral transmission rates (*R*_*t*_=1.2 and *R*_*t*_=1.4). For higher transmission levels, a recent study similarly found that vaccinating high-risk groups first constituted the optimal use of available vaccines [15]. On the other hand, moderate transmission levels are a more realistic scenario as most counties continue to implement moderate social distancing measures during vaccination. Alternative scenarios may be considered for the transmission rate, but the overall outcomes are not expected to be substantially influenced as the current assumptions regarding *R*_*t*_ may be thought of as an “average” version of a time-varying rate. In addition, it is known [1] that the final size of a stochastic epidemic is invariant to the presence of an exposed period and to different distributional assumptions on the infectious period duration. Hence, these assumptions will not alter the conclusions of this work. Other recent relevant results supporting our assumptions include [40,41].

The main conclusions of the present study and all relevant assumptions made about vaccine efficacy are in broad agreement with the results obtained using optimization techniques for a model calibrated using data from Italy [18], despite the fact that the authors only allowed for the vaccine to protect against transmission and not disease. Recent results from Israel suggest that there is also protection against hospitalization and death [35] and therefore these results may be conservative. Similar conclusions are reported when varying vaccine availability and using alternative efficacy assumptions [19]. Assuming that efficacy remains constant after the first dose, [42] used simulation and showed that the effectiveness of vaccination programmes in reducing infections, hospitalizations and deaths is maximized with a delay of 12 to 15 weeks for both the Pfizer and Moderna vaccines. Similar recommendations on delaying the second dose for individuals below the age of 65 are made in [43].

Although we have chosen to primarily emphasize the results of the proposed approach in terms of quantities of interest in public health, additional gains are to be expected in terms of social and economic aspects of public life by offering faster vaccine coverage to the economically active population. An empirical application of the proposed approach is effectively followed in the United Kingdom and Canada, and the outcome seems to be a significantly faster reduction in SARS-CoV-2 circulation.

Overall, our results clearly indicate that, in the presence of a limited vaccine supply, distributing all available doses with a 3-month intermediate time interval could offer important advantages in terms of public health as well as to the wider society and the economy.

## Data Availability

All data and code for this project is available at https://github.com/pbarmpounakis/Evaluating-the-effects-of-vaccine-rollout-policies-in-European-countries-A-simulation-study

## Supporting information

### Supporting information A

Graphical presentation of different scenarios

**S1 Fig.**
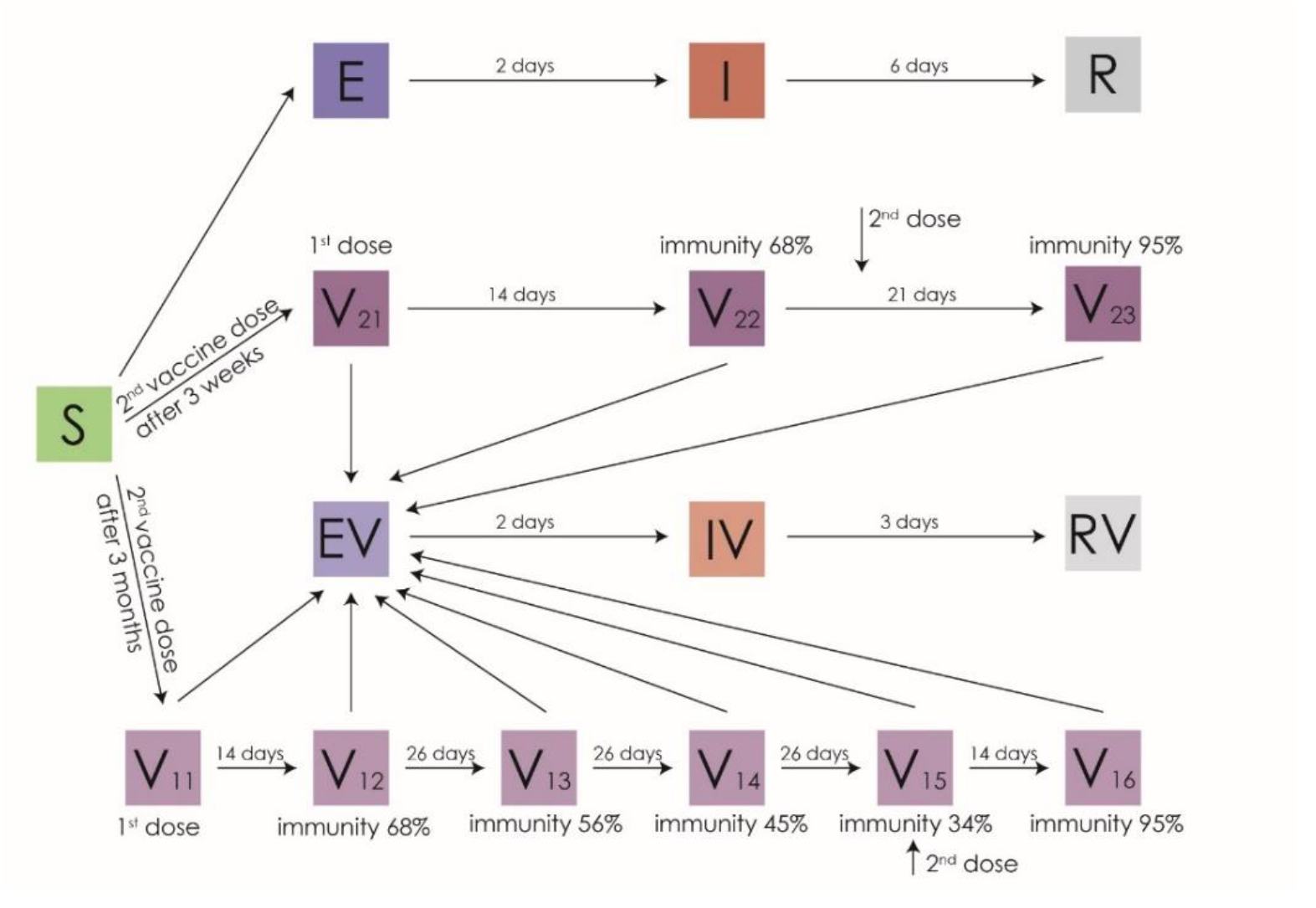
Schematic representation of the S(V)EIR epidemic model for the *worst-case* scenario regarding immunity waning.

**S2 Fig.**
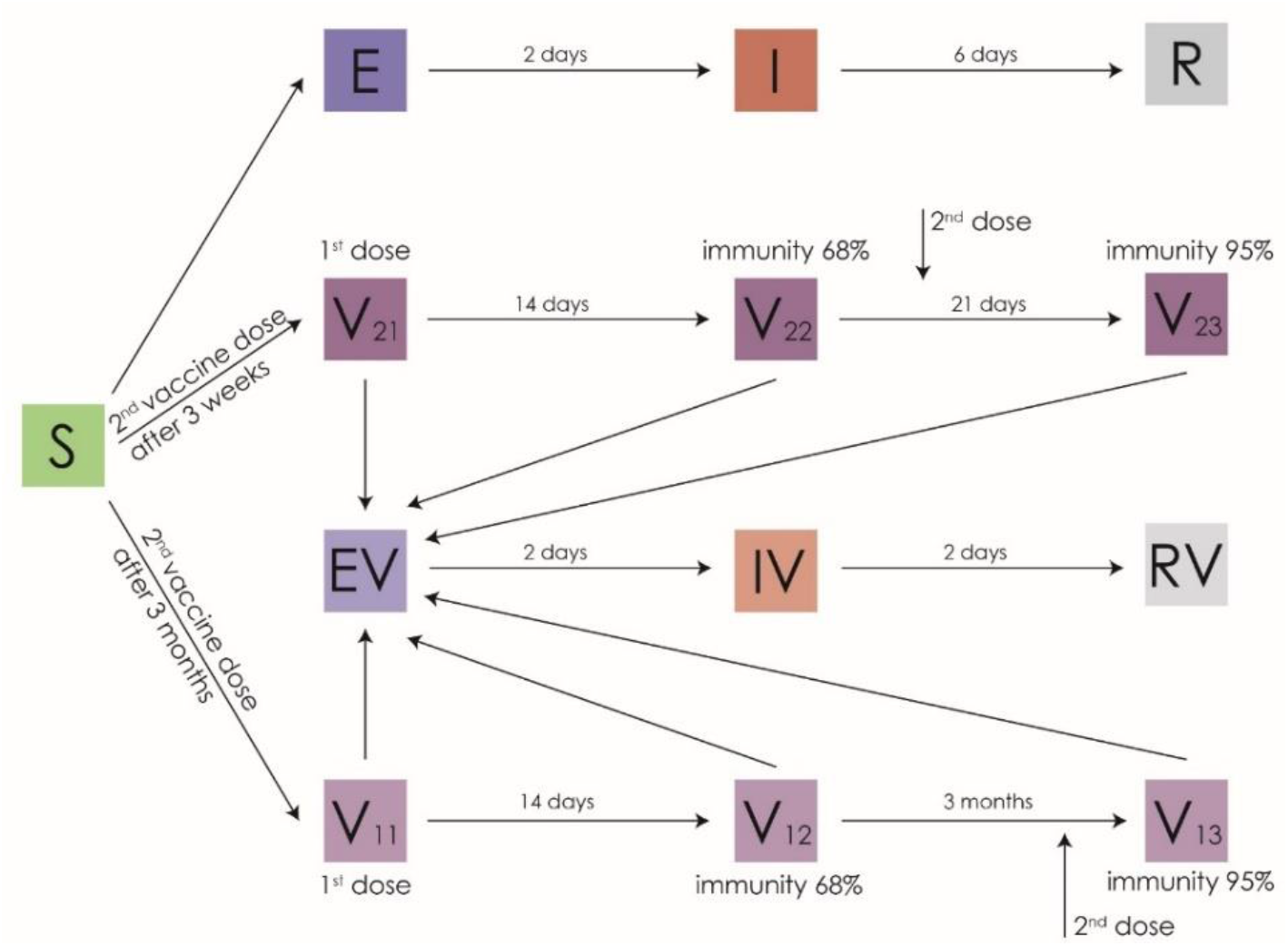
Schematic representation of the S(V)EIR epidemic model for the *optimistic* scenario regarding immunity waning.

### Supporting information B

**S1 Table.**
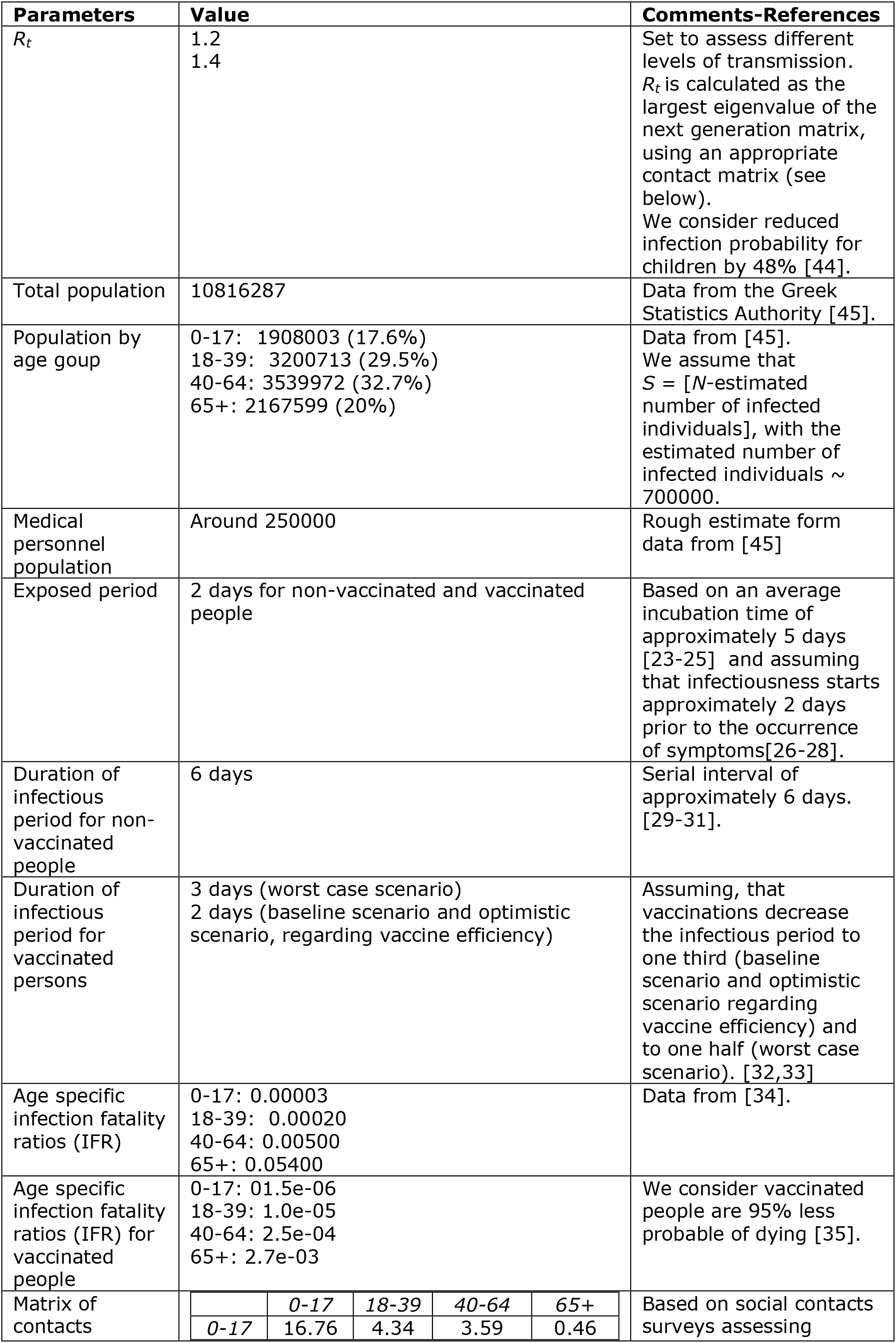
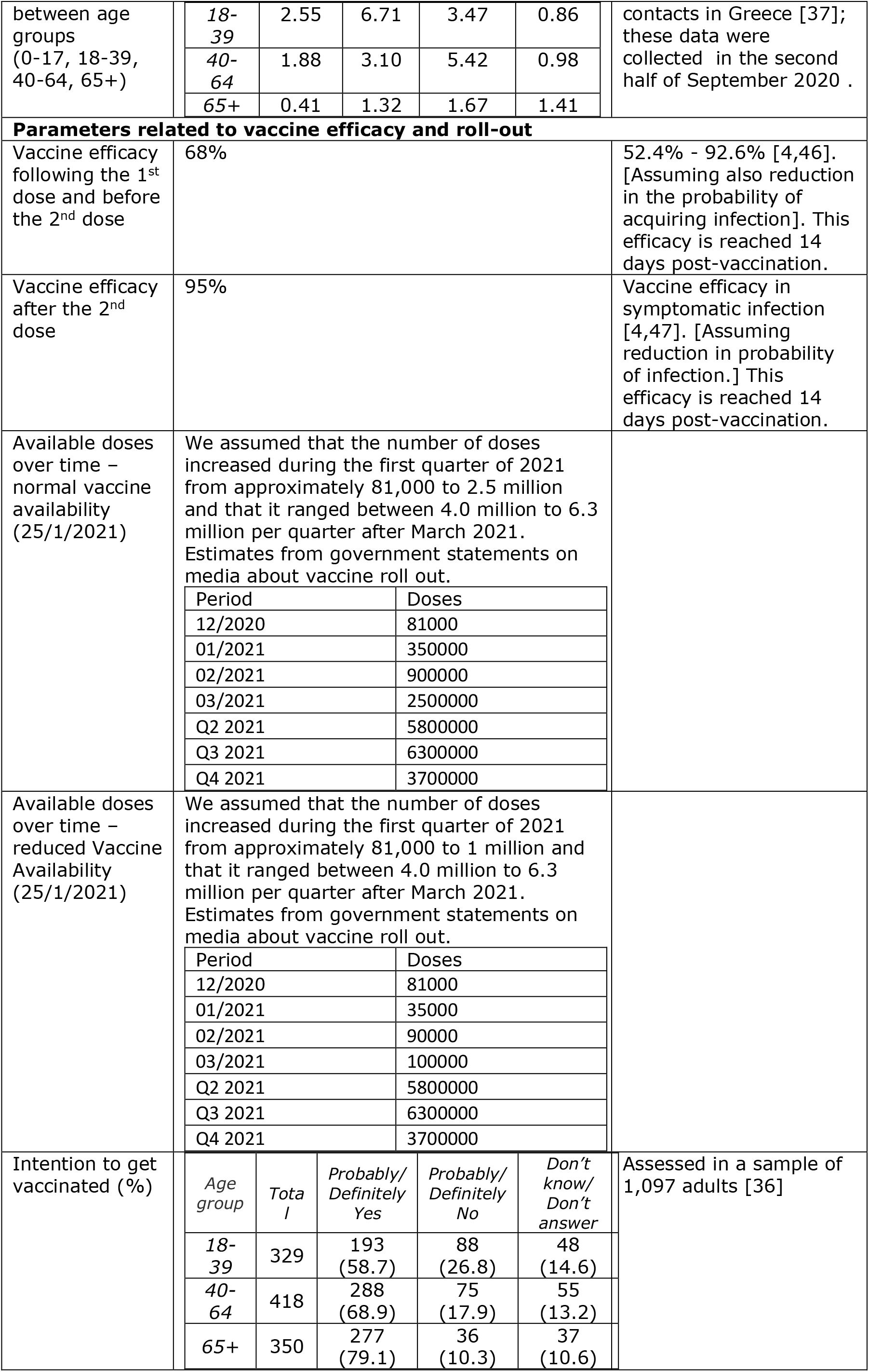
Model Assumptions.

### Supporting information C

Baseline Scenario - Vaccine Availability - *R*_*t*_=1.2

**S2 table.** Cumulative number of deaths, when 0% of vaccines allocated to ages 18-74.

| Cumulative Deaths | End of March | End of June | End of August | End of October | End of December |
| --- | --- | --- | --- | --- | --- |
| 0-17 | 6 (6-6) | 14 (14-14) | 15 (15-15) | 16 (16-16) | 17 (17-17) |
| 18-39 | 75 (74-75) | 143 (143-144) | 146 (145-146) | 146 (146-146) | 146 (146-146) |
| 40-64 | 1854 (1846-1863) | 3649 (3646-3652) | 3747 (3745-3749) | 3748 (3746-3750) | 3749 (3747-3751) |
| 65+ | 1897 (1886-1906) | 2384 (2373-2393) | 2425 (2416-2436) | 2461 (2451-2471) | 2495 (2485-2505) |
| Total deaths | 3832 (3812-3850) | 6190 (6176-6203) | 6333 (6321-6346) | 6371 (6359-6383) | 6407 (6395-6419) |
| Total life years lost | 73352.5 (72982-73685.5) | 134837.5 (134670.5-135044) | 138298.5 (138122-138435.5) | 138654 (138524-138784) | 138995.5 (138865.5-139125.5) |

**S3 Table.** Cumulative number of deaths, when 20% of vaccines allocated to ages 18-74.

| **Cumulative deaths** | End of March | End of June | End of August | End of October | End of December |
| --- | --- | --- | --- | --- | --- |
| 0-17 | 6 (6-6) | 14 (14-14) | 15 (15-15) | 16 (16-16) | 17 (17-17) |
| 18-39 | 75 (74-75) | 139 (139-139) | 141 (141-141) | 141 (141-141) | 141 (141-141) |
| 40-64 | 1852 (1844-1859) | 3550 (3547-3552) | 3632 (3630-3634) | 3634 (3631-3635) | 3635 (3632-3636) |
| 65+ | 1910 (1900-1919) | 2498 (2488-2506) | 2542 (2533-2551) | 2580 (2571-2589) | 2617 (2609-2626) |
| Total deaths | 3843 (3824-3859) | 6201 (6188-6211) | 6330 (6319-6341) | 6371 (6359-6381) | 6410 (6399-6420) |
| Total life years lost | 73383.5 (73020-73656.5) | 132451.5 (132291.5-132567.5) | 135400 (135277-135523) | 135799.5 (135646.5-135892.5) | 136162 (136016-136255) |

**S4 Table.**
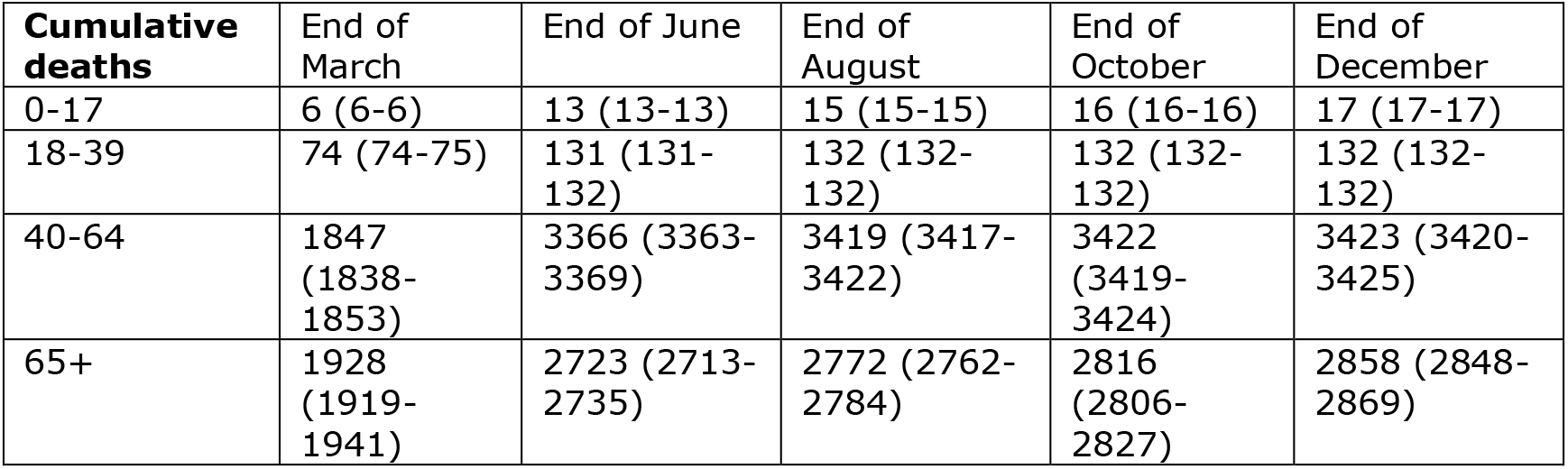
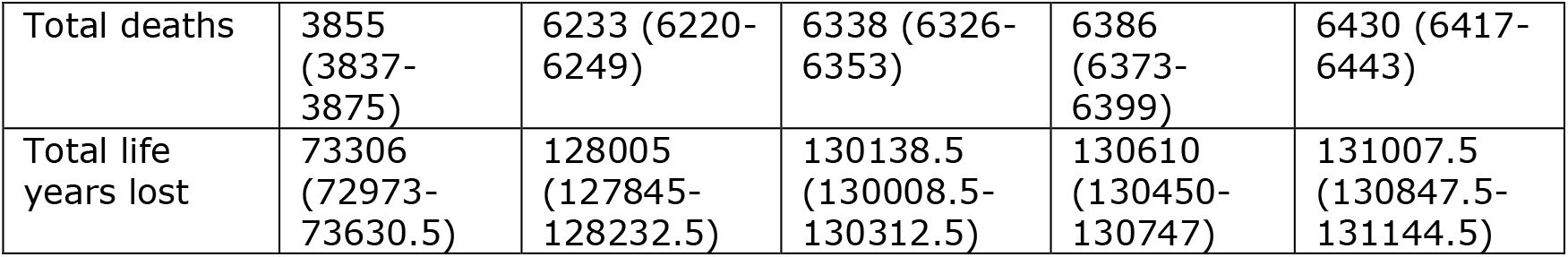
Cumulative number of deaths, when 50% of vaccines allocated to ages 18-74.

| **Cumulative deaths** | End of March | End of June | End of August | End of October | End of December |
| --- | --- | --- | --- | --- | --- |
| 0-17 | 6 (6-6) | 13 (13-13) | 15 (15-15) | 16 (16-16) | 17 (17-17) |
| 18-39 | 74 (74-75) | 131 (131-132) | 132 (132-132) | 132 (132-132) | 132 (132-132) |
| 40-64 | 1847 (1838-1853) | 3366 (3363-3369) | 3419 (3417-3422) | 3422 (3419-3424) | 3423 (3420-3425) |
| 65+ | 1928 (1919-1941) | 2723 (2713-2735) | 2772 (2762-2784) | 2816 (2806-2827) | 2858 (2848-2869) |
| Total deaths | 3855<br>(3837-3875) | 6233 (6220-6249) | 6338 (6326-6353) | 6386 (6373-6399) | 6430 (6417-6443) |
| Total life years lost | 73306<br>(72973-73630.5) | 128005<br>(127845-128232.5) | 130138.5<br>(130008.5-130312.5) | 130610<br>(130450-130747) | 131007.5<br>(130847.5-131144.5) |

**S5 Table.** Cumulative number of deaths, when 100% of vaccines allocated to ages 18-74.

| **Cumulative deaths** | End of March | End of June | End of August | End of October | End of December |
| --- | --- | --- | --- | --- | --- |
| 0-17 | 6 (6-6) | 13 (13-13) | 15 (15-15) | 16 (16-16) | 17 (17-17) |
| 18-39 | 75 (74-75) | 129 (129-130) | 130 (130-130) | 130 (130-130) | 130 (130-130) |
| 40-64 | 1854<br>(1846-1862) | 3304 (3300-3309) | 3311 (3307-3316) | 3313 (3310-3319) | 3314 (3311-3319) |
| 65+ | 1881<br>(1870-1889) | 2278 (2268-2286) | 2312 (2302-2320) | 2340 (2330-2348) | 2367 (2357-2375) |
| Total deaths | 3816<br>(3796-3832) | 5724 (5710-5738) | 5768 (5754-5781) | 5799 (5786-5813) | 5828 (5815-5841) |
| Total life years lost | 73240.5<br>(72870-73536.5) | 122923<br>(122733-123182.5) | 123571.5<br>(123381.5-123777.5) | 123901<br>(123741-124137) | 124193.5<br>(124033.5-124399.5) |

**S6 Table.** Cumulative number of infections, when 0% of vaccines allocated to ages 18-74.

| **Cumulative infections** | End of March | End of June | End of August | End of October | End of December |
| --- | --- | --- | --- | --- | --- |
| 0-17 | 205692<br>(201056-210308) | 407403<br>(395412-419507) | 449126<br>(434390-464064) | 483022<br>(465868-500426) | 514726<br>(495223-534530) |
| 18-39 | 355723<br>(349647-361850) | 657034<br>(642333-671954) | 675324<br>(658976-691931) | 678502<br>(661464-695861) | 681604<br>(663874-699700) |
| 40-64 | 354386<br>(348331-360467) | 668880<br>(654066-683859) | 695936<br>(679082-713033) | 699982<br>(682338-717904) | 703266<br>(684908-721941) |
| 65+ | 42180<br>(40320-44094) | 57708<br>(54237-61357) | 59220<br>(55278-63394) | 60525<br>(56142-65195) | 61809<br>(56997-66971) |

**S7 Table.** Cumulative number of infections, when 20% of vaccines allocated to ages 18-74.

| **Cumulative infections** | End of March | End of June | End of August | End of October | End of December |
| --- | --- | --- | --- | --- | --- |
| 0-17 | 205563<br>(201024-210191) | 403485<br>(391605-415485) | 445046<br>(430414-459859) | 479322<br>(462237-496643) | 511262<br>(491857-531012) |
| 18-39 | 355280<br>(349268-361332) | 643554<br>(628986-658220) | 662802<br>(646528-679252) | 668244<br>(651052-685670) | 671562<br>(653659-689750) |
| 40-64 | 353940<br>(347968-360029) | 655699<br>(641042-670620) | 682914<br>(666185-699958) | 689238<br>(671522-707328) | 693104<br>(674617-712012) |
| 65+ | 42170<br>(40289-44097) | 58765<br>(55217-62461) | 60310<br>(56284-64541) | 61671<br>(57194-66410) | 63013<br>(58098-68248) |

**S8 Table.** Cumulative number of infections, when 50% of vaccines allocated to ages 18-74.

| **Cumulative infections** | End of March | End of June | End of August | End of October | End of December |
| --- | --- | --- | --- | --- | --- |
| 0-17 | 205382<br>(200846-210114) | 396362<br>(384695-408329) | 437560<br>(423185-452361) | 472510<br>(455670-489830) | 504882<br>(485694-524631) |
| 18-39 | 354592<br>(348511-360648) | 618118<br>(603982-632366) | 638982<br>(623014-655121) | 646856<br>(629797-664156) | 650152<br>(632378-668213) |
| 40-64 | 353262<br>(347216-359440) | 630735<br>(616477-645284) | 658194<br>(641843-674894) | 667522<br>(649974-685486) | 671722<br>(653378-690546) |
| 65+ | 42128<br>(40257-44041) | 60744<br>(57067-64557) | 62360<br>(58188-66723) | 63826<br>(59184-68701) | 65248<br>(60157-70642) |

**S9 Table.** Cumulative number of infections, when 100% of vaccines allocated to ages 18-74.

| **Cumulative infections** | End of March | End of June | End of August | End of October | End of December |
| --- | --- | --- | --- | --- | --- |
| 0-17 | 205698<br>(201113-210283) | 394808<br>(383203-406569) | 434849<br>(420571-449360) | 470048<br>(453294-487112) | 502502<br>(483371-521985) |
| 18-39 | 355640<br>(349658-361635) | 611915<br>(598340-625688) | 638422<br>(622750-654357) | 645310<br>(628653-662313) | 648467<br>(631115-666220) |
| 40-64 | 354362<br>(348317-360332) | 623402<br>(609535-637325) | 651674<br>(635668-667804) | 661004<br>(643853-678331) | 664376<br>(646514-682482) |
| 65+ | 42111<br>(40235-44022) | 59390<br>(55683-63234) | 61865<br>(57562-66355) | 63053<br>(58343-68022) | 64212<br>(59091-69658) |

